# Probing Human Heart TCA Cycle Metabolism and Response to Glucose Load using Hyperpolarized [2-^13^C]Pyruvate MR Spectroscopy

**DOI:** 10.1101/2023.10.16.23297053

**Authors:** Hsin-Yu Chen, Jeremy W. Gordon, Nicholas Dwork, Brian T. Chung, Andrew Riselli, Sanjay Sivalokanathan, Robert A. Bok, James B. Slater, Daniel B. Vigneron, M. Roselle Abraham, Peder E.Z. Larson

## Abstract

**Introduction:** The normal heart has remarkable metabolic flexibility that permits rapid switching between mitochondrial glucose oxidation and fatty acid (FA) oxidation to generate ATP. Loss of metabolic flexibility has been implicated in the genesis of contractile dysfunction seen in cardiomyopathy. Metabolic flexibility has been imaged in experimental models, using hyperpolarized (HP) [2-^13^C]pyruvate MRI, which enables interrogation of metabolites that reflect tricarboxylic acid (TCA) cycle flux in cardiac myocytes. This study aimed to develop methods, demonstrate feasibility for [2-^13^C]pyruvate MRI in the human heart for the first time, and assess cardiac metabolic flexibility.

**Methods:** Good Manufacturing Practice [2-^13^C]pyruvic acid was polarized in a 5T polarizer for 2.5-3 hours. Following dissolution, QC parameters of HP pyruvate met all safety and sterility criteria for pharmacy release, prior to administration to study subjects. Three healthy subjects each received two HP injections and MR scans, first under fasting conditions, followed by oral glucose load. A 5cm axial slab-selective spectroscopy approach was prescribed over the left ventricle and acquired at 3s intervals on a 3T clinical MRI scanner.

**Results:** The study protocol which included HP substrate injection, MR scanning and oral glucose load, was performed safely without adverse events. Key downstream metabolites of [2-^13^C]pyruvate metabolism in cardiac myocytes include the glycolytic derivative [2-^13^C]lactate, TCA-associated metabolite [5-^13^C]glutamate, and [1-^13^C]acetylcarnitine, catalyzed by carnitine acetyltransferase (CAT). After glucose load, ^13^C-labeling of lactate, glutamate, and acetylcarnitine from ^13^C-pyruvate increased by 39.3%, 29.5%, and 114%, respectively in the three subjects, that could result from increases in lactate dehydrogenase (LDH), pyruvate dehydrogenase (PDH), and CAT enzyme activity as well as TCA cycle flux (glucose oxidation).

**Conclusions:** HP [2-^13^C]pyruvate imaging is safe and permits non-invasive assessment of TCA cycle intermediates and the acetyl buffer, acetylcarnitine, which is not possible using HP [1-^13^C]pyruvate. Cardiac metabolite measurement in the fasting/fed states provides information on cardiac metabolic flexibility and the acetylcarnitine pool.

**Graphical Abstract:** 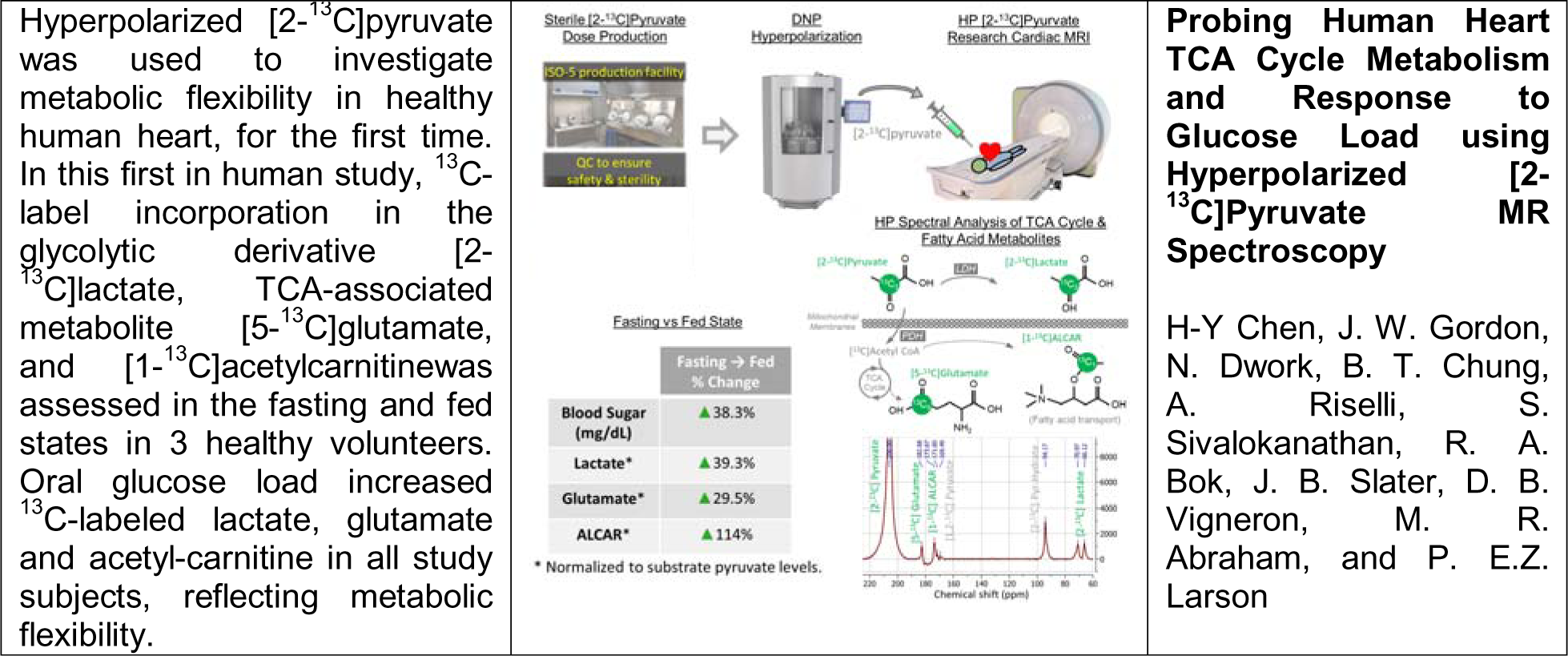

## 1. Introduction

Heart disease is the leading cause of death both globally and in the US^1^. A disturbance of cardiac energy metabolism is an early feature of heart disease because of the high energy demands imposed by cardiac contraction. Mitochondria are the cellular powerhouses of cardiac myocytes with mitochondrial oxidative metabolism generating approximately 95% of ATP, primarily through an interconnected network of complex metabolic machinery involving the tricarboxylic acid (TCA) cycle and oxidative phosphorylation^2,3^. Changes in substrate and oxygen availability, cardiac work, mitochondrial function and mutant sarcomeric proteins^4^ cause secondary changes in gene expression and structural remodeling^5^, that predispose to heart failure and arrhythmias.

While assessing structure and function of the heart can be done clinically via multiple modalities (Echo, CT, MRI), the capabilities for cardiac metabolic imaging are limited^6–8^. Cardiac imaging with positron emission tomography (PET) has been readily applied to investigate glucose and fatty acid (FA) uptake in the heart using radiotracers such as FDG, ^11^C-glucose, ^11^C-acetate, and ^11^C-palmitate^5,9,10^. Nevertheless, PET does not follow the metabolic fate of these radiolabeled probes beyond their initial distribution and uptake, nor does it distinguish the chemical species of the downstream products from the injected substrates ^11,12^. ^31^P and ^1^H MR spectroscopy have shown the ability to measure energetics (ATP and creatine) and metabolite levels, respectively, but require long scan times of > 10 minutes and have low spatial resolution (>10cc voxels). Metabolic imaging with Deuterium-labelled compounds showed some initial promises in animal models, but suffers from similar limitations in sensitivity^13,14^.

Hyperpolarized (HP) ^13^C MRI takes advantage of safe, non-radioactive ^13^C isotope-labelled agents and dynamic nuclear polarization technology to provide unprecedented 50,000-fold sensitivity enhancement to offer real-time measurements indicative of metabolic activity in humans, including glycolytic metabolism and blood flow as a rapid, 2-minute addition to a regular clinical MRI exam^11,15–20^. The distinct chemical shifts between injected HP substrate and each downstream product not only allow identification of each chemical species of interest, but provides direct measurement of metabolic activity. Initial imaging studies of the human heart using HP ^13^C pyruvate, enriched at C1 position ([1-^13^C]pyruvate), detected metabolic conversion in the myocardium to [1-^13^C]lactate, catalyzed by lactate dehydrogenase (LDH), and [^13^C]bicarbonate, catalyzed by pyruvate dehydrogenase (PDH) ^21^. Subsequent research in patients with type 2 diabetes mellitus (T2DM) and healthy volunteers found significantly decreased pyruvate to bicarbonate conversion in T2DM patients, suggestive of impaired cardiac energetics secondary to metabolic disorders^22^. Other cardiac HP [1-^13^C]pyruvate studies explored cardiac metabolism in patients following adenosine infusion for cardiac stress testing, after myocardial infarction and cardiotoxic chemotherapy for breast cancer treatment ^19,23,24^.

The TCA cycle is the central hub of oxidative metabolism, and is dysregulated in mitochondrial dysfunction and cardiomyopathies ^25–27^. A limitation of HP [1-^13^C]pyruvate MRI is that it cannot measure metabolic dynamics indicative of TCA cycle flux, since entry into the TCA cycle via PDH directs the carbon-13 label to ^13^CO_2_, which is then exchanged with [^13^C]bicarbonate. Pyruvate enriched in the C2 position ([2-^13^C]pyruvate; **Figure 1**), on the other hand, passes the carbon-13 label into the mitochondrial TCA cycle through acetyl-CoA ^28–30^, which allows novel additional measurements of the downstream TCA-associated intermediates such as [5-^13^C]glutamate and [5-^13^C]glutamine/[1-^13^C]citrate. Furthermore, HP [2-^13^C]pyruvate can also be converted into [1-^13^C]acetylcarnitine (ALCAR), mediated first by PDH to acetyl-CoA and then by carnitine acetyltransferase (CAT). Acetylcarnitine serves as an acetyl CoA buffer^12^ and fine tunes myocyte availability of acetyl-CoA that fuels the TCA cycle and oxidative phosphorylation ^3,12,31–33^. Acetylcarnitine levels also reflect myocytic stores of carnitine, which transports long chain FA into mitochondria via the carnitine shuttle. A few other HP ^13^C probes, such as acetate^34^, octanoate^35^, butyrate^36^, and Acetyl-L-Carnitine^37^ are also of substantial interest for TCA cycle metabolic investigations, but these compounds are yet to be translated for human studies.

**Figure 1.**
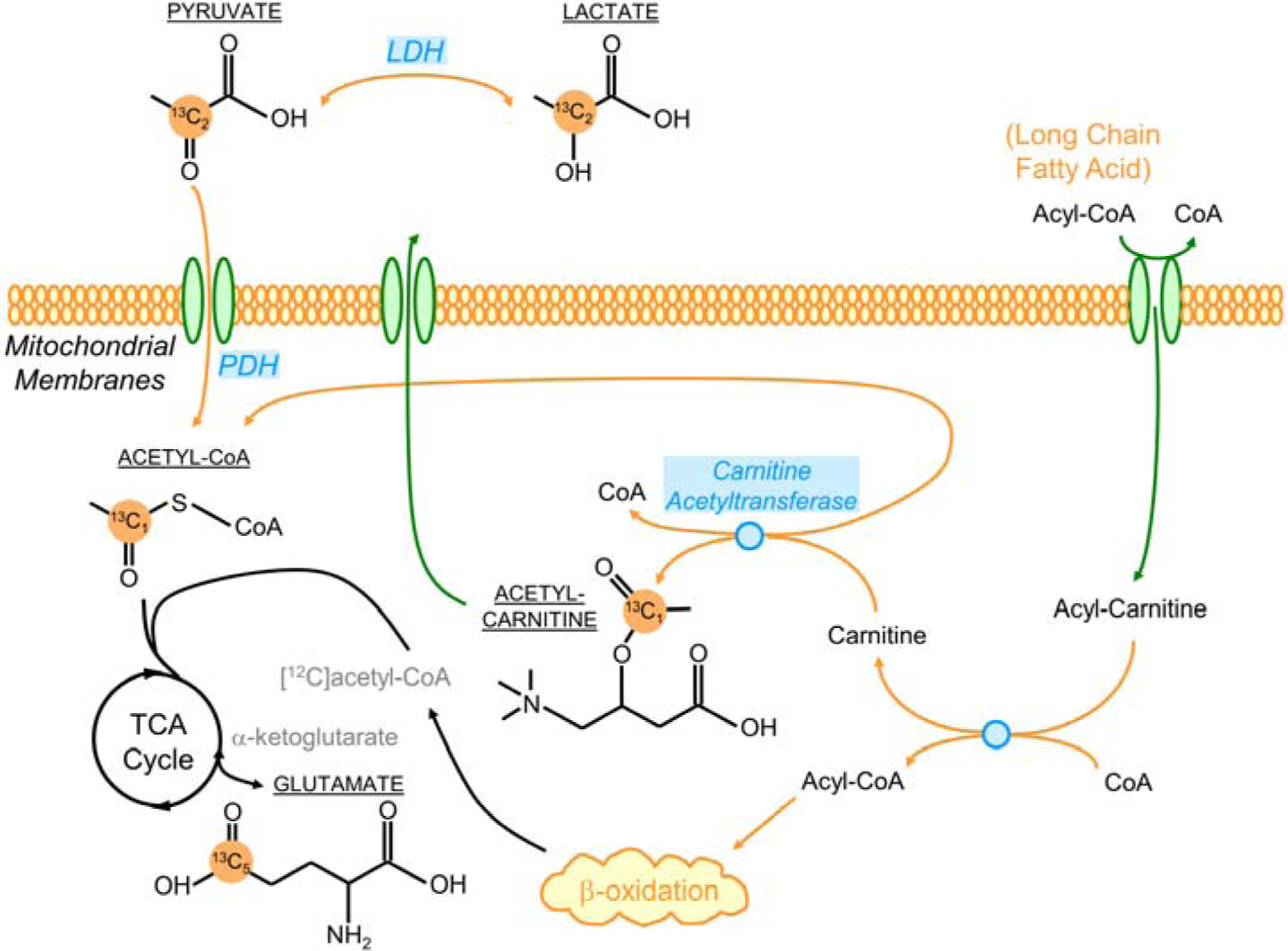
A diagram depicting the flux of [2-^13^C]pyruvate label into the TCA cycle and the acetylcarnitine, showing the propagation of C-13 labeling in this study. Acetylcarnitine not only serves as an acetyl CoA buffer, but is involved in the “carnitine shuttle” which transports long chain fatty acid into mitochondria for β oxidation.

The objective of this pilot study was to develop methods, examine the safety and feasibility of hyperpolarized [2-^13^C]pyruvate MRI, and assess metabolic flexibility in human myocardium, for the first time. We used pharmaceutical preparation methods to create human HP [2-^13^C]pyruvate doses and developed a specialized MRI pulse sequence to collect the data after injection. To modulate substrate availability, HP scans were acquired under fasting conditions and 30 minutes after oral glucose administration in healthy volunteers. Outcome measures included pharmaceutical production success, QC release and delivery of the HP agent, safety and tolerability of the agent, and finally the ability to detect, identify and quantify derivatives of HP [2-^13^C]pyruvate, namely [5-^13^C]glutamate, [2-^13^C]lactate and [1-^13^C]acetylcarnitine. These studies offer insights into cardiac energy metabolism, metabolic flexibility, mitochondrial function and enable development of metabolic therapies for heart disease.

## 2. Methods

### Subject Characteristics

Three healthy volunteer subjects were enrolled through an IRB-approved protocol (UCSF IRB 19-27889). All subjects were healthy males, age 33-42 years (median: 41 y/o).

Subject demographics, vitals and a basic metabolic panel was obtained (**Table 1A**). Physical fitness of study subjects was evaluated using an abbreviated version of the standardized Minnesota Leisure Time Physical Activities questionnaire^38^. The questionnaire assessed the type, frequency, duration and intensity of physical activities on a weekly and monthly basis around the timeframe of the MRI study.

**Table 1.** **A)** Summary of patient demographics, basic metabolic panel and an exercise questionnaire. All laboratory results were within the normal range without significant readings that may indicate any cardiovascular or metabolic conditions. Results from the Minnesota questionnaire showed two subjects could be categorized as physically active, whereas one was physically inactive. **B)** Basic evaluation of cardiac anatomy based on 1H MRI showed no anomalies.

| A) | Age (years) | Race | Gender | BMI | Blood Pressure | Heart Rate (bpm) | Insulin (mU/L) | Ketones (mmol/L) | Fatty Acids (mg/dL) | Medications |
| --- | --- | --- | --- | --- | --- | --- | --- | --- | --- | --- |
| Subject 1 | 38 ± 5 | White | Male | 24.2 | 123/71 | 64 | 4.6 | 0.10 | NA | None (x2)<br>Yes (x1)* |
| Subject 2 |  | White | Male | 25.3 | 116/60 | 47 | 5.7 | 0.31 | 94 |  |
| Subject 3 |  | Asian/Pacific Islander | Male | 23.1 | NA | 75 | 5.9 | 0.12 | 47 |  |

\* One subject reported use of prescription medication unrelated to cardiovascular or metabolic conditions.

|  | Self-reported activity | Years physically active | Types of activities | Days/wk physically active | Hours/wk physically active | Min/day walking | Days/week walking | Types of home activities |
| --- | --- | --- | --- | --- | --- | --- | --- | --- |
| Subject 1 | Active | > 6 | Cycling, swimming, hiking | 2-3 | 4-5 | 10-30 | 3-4 | Cooking, taking out trash, cleaning |
| Subject 2 | Inactive | 0-2 | N/A | 0-1 | 0-1 | 10-30 | 3-4 | *Not answered |
| Subject 3 | Active | > 6 | Running, hiking | 2-3 | 2-3 | 10-30 | >5 | cooking, carrying out trash, laundry, carrying groceries upstairs, cleaning bathroom |

**Table 1. A)** Summary of patient demographics, basic metabolic panel and an exercise questionnaire.
| B) | LV end-diastolic diameter (mm) | LV end-diastolic volume (ml) | RV end-diastolic volume (ml) | LV wall thickness (mm) | LA longitudinal diameter (mm) | LA transverse diameter (mm) | LA volume (ml) | RA longitudinal diameter (mm) | RA transverse diameter (mm) | RA volume (ml) |
| --- | --- | --- | --- | --- | --- | --- | --- | --- | --- | --- |
| Subject 1 | 53 | 197 | 91 | 5.7 | 2.9 | 2.4 | NA* | 3.5 | 4.9 | 81 |
| Subject 2 | 54 | 168 | 108 | 5.5 | 3.8 | 4.1 | 48.7 | 3.3 | 3.9 | 46 |
| Subject 3 | 52 | 192 | NA* | 6.5 | 3.6 | 3.5 | 51.2 | 4.3 | 4.3 | 63 |

\* Data excluded due to insufficient image quality for evaluation.

### Hyperpolarized ^13^C MRI exam

Pharmacy kits containing Good Manufacturing Practice (GMP)-grade [2-^13^C]pyruvic acid (Isotec, MilliporeSigma, Burlington MA) were manufactured and polarized in a 5T clinical polarizer (SPINLab, GE Healthcare, Chicago IL) for 2.5-3 hours following a published workflow ^28^. The hyperpolarized [2-^13^C]pyruvate was then rapidly dissoluted, neutralized, and underwent terminal sterilization. Pharmacy release followed confirmation that all QC parameters satisfied safety and sterility specifications, including pyruvate and EPA concentrations, polarization percentage, pH, temperature, and bubble point test of the sterility filter. The ^13^C scans were initiated 5 seconds after the start of injection. The MRI exams were conducted on a clinical 3T MRI scanner (MR750, GE Healthcare, Chicago IL) equipped with multinuclear spectroscopy capability. The anatomical imaging used the body coil for transmit, and signal reception used either a specialized 4-channel torso array receiver, or a commercially available flexible 21-channel array receiver (AIR Coil, GE Healthcare, Chicago IL). The hardware setup for hyperpolarized ^13^C imaging included a clamshell Helmholtz transmitter as previously described, and a specialized rigid array consisting of two 4-element receiver “paddles” for signal reception ^39^. As shown in **Figure 2**, the two paddles were positioned anteriorly and laterally over the left chest wall, respectively. Anatomical reference images of the heart were acquired using an axial short TE gradient echo (TE = 1.08ms, FOV = 35cm, in plane resolution = 0.21mm, slice thickness = 3mm) and a coronal balanced steady-state free precession sequence (TE/TR = 1.36ms/3.08ms, FOV = 36cm, in plane resolution = 0.24mm, slice thickness = 5mm). The former is applied to pick up the built-in Vitamin E fiducial markers on the paddle receivers to ensure appropriate coil positioning.

**Figure 2.**
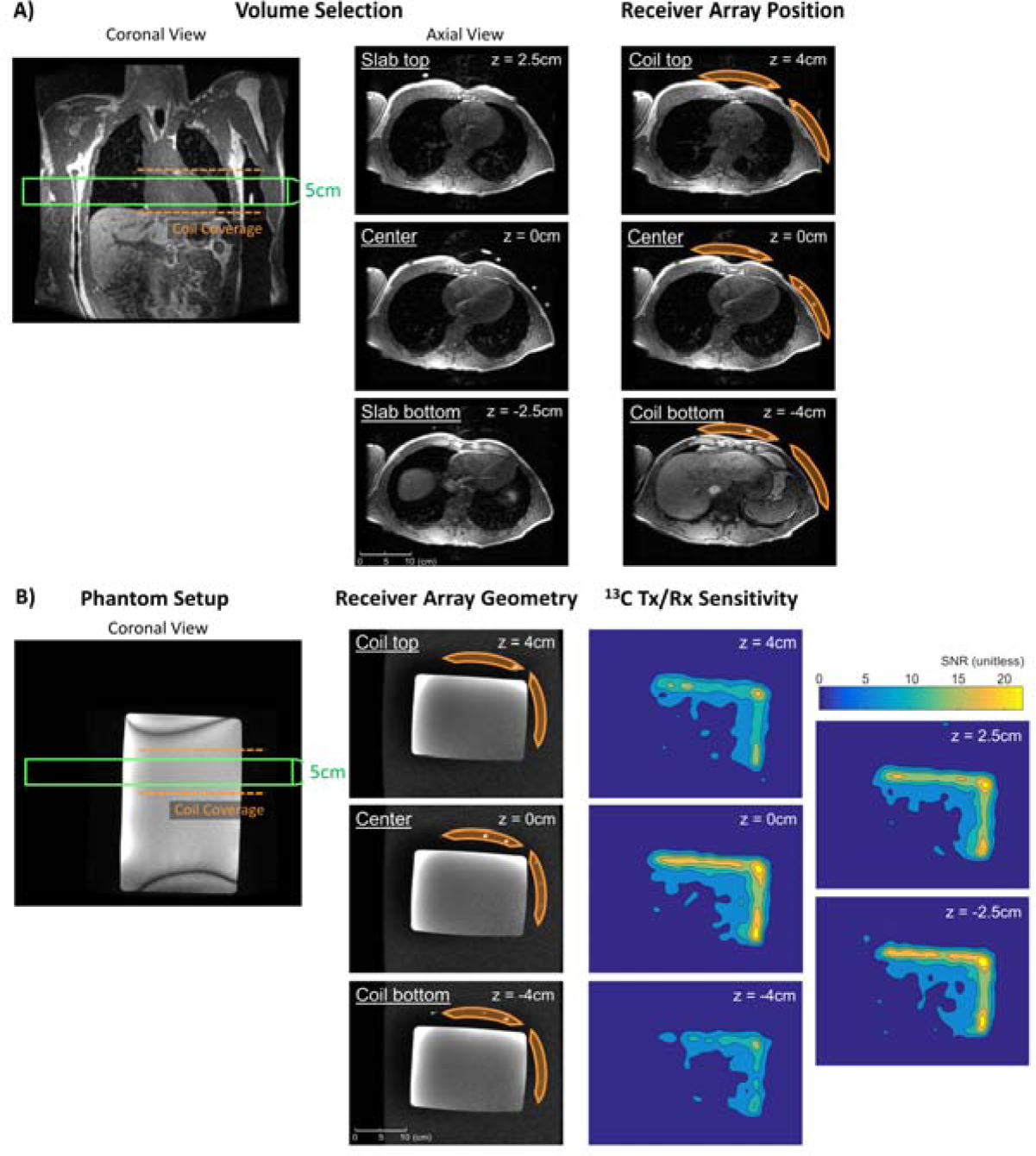
Volunteer and phantom setup. **A)** For volunteer studies, the two paddle receivers were placed anteriorly and laterally over the subject’s left chest wall, respectively. Based off the ^1^H anatomical reference images, the axial 5cm RF excitation slice was prescribed to cover the left ventricle, shown as the green box. **B)** A measured approximate sensitivity profile accounting for both the B1^+^ field of the clamshell transmitter and the B1^-^ field of the paddle receiver arrays. The receivers were arranged anterior and lateral to a rectangular dimethyl silicon phantom to emulate the setup for human studies. Each paddle coil covered a ∼10×8cm area, and detected ∼8cm depth in its direct line of sight. Factoring in the anatomical structures covered by the intersection the Tx/Rx sensitivity and the RF excitation profiles, the signal would have predominantly come from the heart. This configuration provided basic spatial localization despite the spectroscopic nature of our acquisition approach.

### Oral Glucose Load Test and Measurement

The study subjects were instructed to undergo at least 4 hours of fasting prior to the baseline hyperpolarized pyruvate scan. After the baseline scan, the subjects were given 350mL of sugar drink containing approximately 29 grams of sugar (dextrose, sucrose; Gatorade, Chicago IL) for oral ingestion^40^. A post-glucose hyperpolarized scan was acquired ∼30 minutes after the oral glucose load.

The blood glucose readings at baseline fasting and after glucose load were performed using a commercially available glucometer (Contour Next Glycometer, Ascensia, Switzerland) in a standardized fashion. Blood samples were collected from the IV access site at the antecubital vein and applied on test strips to obtain readings. Per protocol, the readings were taken right before each hyperpolarized ^13^C injection/scan.

### ^13^C Pulse Sequence

Dynamic slab spectroscopy (TE = 3.7ms) was acquired at 3s intervals over a 90s timeframe, targeting a 5cm-thick axial slab prescribed over the left ventricle. The acquisition bandwidth was 8000Hz, acquisition time was ∼256 ms, and the spectral resolution was ∼3.9Hz.

The relatively wide chemical shift range (∼150ppm) among [2-^13^C]pyruvate and its downstream products poses certain challenges for selectively exciting each species with regards to both spatial localization and metabolite-specific flip angles. To this end, a specialized broadband spectral-spatial RF pulse was designed for the excitation of [2-^13^C]pyruvate and its downstream products in the human heart with metabolite-specific flip angles (**Figure 3**). The substrate [2-^13^C]pyruvate was excited with a 5° tip angle, whereas the products including [5-^13^C]glutamate, [5-^13^C]glutamine/[1-^13^C]citrate, [1-^13^C]acetylcarnitine (ALCAR), and [2-^13^C]lactate each were excited with a 20° tip angle. A 12ppm passband with 1% ripple allowance covered the main TCA cycle-associated metabolites (glutamate, glutamine/citrate, ALCAR), and a 2ppm and a 4ppm passband with 0.2% ripple each accounted for pyruvate and lactate resonances. A spectral correction technique^41^ was applied to eliminate the slice displacement that normally occur due to chemical shift. The time-bandwidth product was set to 3 to fully utilize the gradient and slew rate performance. The RF pulse was designed using an open-source hyperpolarized-MRI-toolbox^42^.

**Figure 3.**
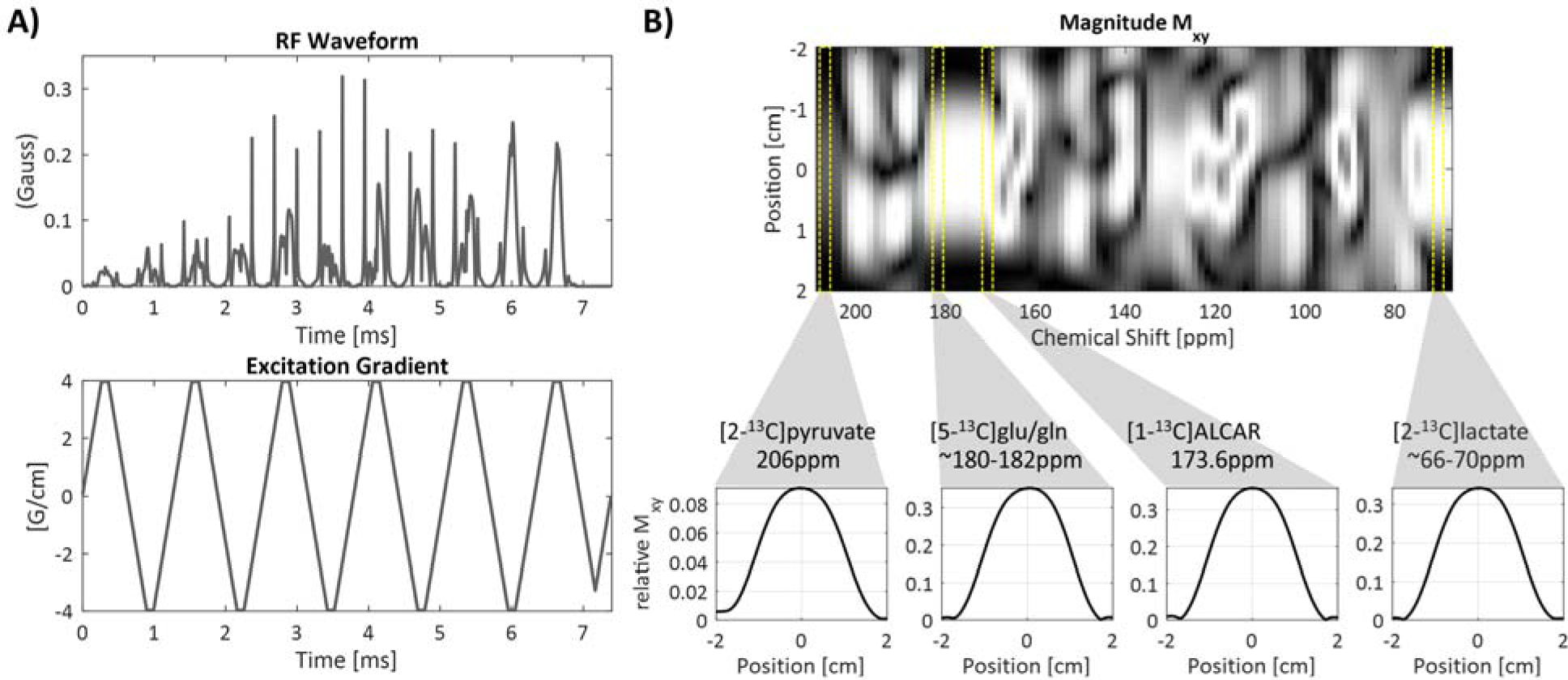
A specialized broadband spectral-spatial RF design enabled excitation of [2-^13^C]pyruvate substrate and its downstream products including [2-^13^C]lactate, [5-^13^C]glutamate/glutamine(or [1-^13^C]citrate), and [1-^13^C]acetylcarnitine (ALCAR) across an approximately 150ppm range. **A)** The 6.4ms broadband pulse utilizes symmetric EPI gradient for improved spatial profile, where time-bandwidth was set to 3 to fully utilize the gradient and slew rate performances. **B)** Spectral-spatial excitation profile shows desirable spatial selectivity at the resonance frequency of each metabolite, where [2-^13^C]pyruvate is excited with a 5° flip, and [2-^13^C]lactate, [5-^13^C]glu/gln, and [1-^13^C]ALCAR each sees a 20° flip. Spectral correction eliminated the chemical shift slice offsets, ensuring the spatial profiles of all resonances of interest were centrally aligned as shown in the individual metabolite profiles.

Center frequency for the HP ^13^C acquisition was determined using a standardized conversion factor from the automatically-calibrated proton frequency as previously described^43^. A fixed offset of 193Hz was added to account for the delta frequency between [1-^13^C]pyruvate calculated by the conversion factor and the center frequency specified in the spectral-spatial RF pulse design (which was 193Hz downfield to [1-^13^C]pyruvate). The transmit power was calibrated using a glass head phantom containing ethylene glycol.

### Data Processing and Analysis

The time-resolved ^13^C spectroscopy data was first combined using a phase-sensitive, optimally weighted sum across receiver channels, and fed through a signal enhancement (synonymously, denoising) filter in MATLAB (Mathworks, Natick MA) using a simple SVD algorithm with rank 10 ^44^, which accounts for the number of substrate and product chemical species as well as nuisance resonances. Subsequently, a mild line broadening of 5Hz Gaussian and a baseline correction using the spline algorithm were applied on MestreNova (Mestrelab Research, Santiago de Compostela, Spain). Quantitative measurement of the dynamic HP profile was calculated by integrating the area under the ppm range of each resonance, and summing associated areas for any multiplet chemical species. Each species was individually phased for quantification. The area-under-the-curve(AUC) spectra for display were calculated by summing the spectrum at each individual timeframe through time. Metabolic conversion was quantified using ratiometric measures of products divided by pyruvate, summed over time.

## 3. Results

### Safety and Technical Feasibility

The administration of C2-labelled HP pyruvate, the MRI scans and the oral glucose load test were safe and well-tolerated without adverse events in the three volunteers enrolled in this study, each of whom received 2 doses of HP [2-^13^C]pyruvate injection with an approximate 30-minute interval between doses. The safety profile is in good agreement with published HP ^13^C studies of cardiac metabolism using C1-labelled pyruvate in patients with ischemic heart disease, diabetes, malignancies, as well as healthy volunteers ^19,21–24,45,46^. The six HP dissolutions yielded 198-245mM (median: 229mM) pyruvate with 27.1-31.2% (median:29.3%) polarization, 0.6-2.2µM (median: 1.6µM) residual trityl radical, pH of 7.5-8.2 (median: 7.8), and 30.7-33.7°C (median: 32.8°C) temperature.

### Subject Demographics and Physical Fitness

The age of the three subjects was 38 ± 5 years. The median BMI was 24.2 (range: 23.1-25.3), where two individuals fell within the healthy weight range and one was borderline overweight based on CDC guidelines^47^. The reported blood pressure, heart rate, insulin and ketone levels were also within the normal range. Two subjects denied use of prescription medication, whereas one subject reported use of a prescription medication unrelated to and not indicated for cardiovascular or metabolic conditions. Unfortunately, two entries on **Table 1A** were not documented, marked as “NA”.

Since exercise training can influence cardiac energy metabolism and carnitine stores, physical activity was quantified using the Minnesota questionnaire. Two subjects were categorized as physically active, whereas one was physically inactive (**Table 1B**). The average number of days per week each subject undertook physical activity ranged from 0-3, and number of hours weekly ranged from 0-5. All subjects reported walking for 10-30 minutes at a time, anywhere from 3 to more than 5 days weekly. All subjects also reported participating in varying amounts and intensities of home activities, such as cooking, cleaning, and taking out trash.

Basic anatomical parameters of the heart were extracted from ^1^H MRI, summarized in **Table 1C**.

### Selectivity of ^13^C Experiment

The slab-selective excitation was placed in the axial plane covering most of the left and right ventricles (**Figure 2A**). The other tissues present in the slab were the skeletal muscle and subcutaneous fat surrounding the chest, the spine, the rib cage, and part of the arms. Of these tissues, only HP pyruvate and downstream metabolites are expected to be observed in the heart and vasculature based on prior HP [1-^13^C]pyruvate imaging results where little to no signal was observed in skeletal muscle and subcutaneous fat ^19,21,23,24^. The placement and sensitivity profile of the receiver coils further improved the localization of HP signals to the heart (**Figure 2B**) and reduced expected major vasculature signal contributions.

### Identification and Quantification of Product Species from [2-^13^C]pyruvate

Setting the substrate [2-^13^C]pyruvate resonance to 206ppm as the reference, the products were presumptively assigned as [5-^13^C]glutamate (182.5ppm), [1-^13^C]acetylcarnitine (ALCAR, 173.6ppm), [2-^13^C]lactate (70.8, 66.2ppm), [1,2-^13^C]pyruvate (171.8, 169.7ppm), and [2-^13^C]pyruvate hydrate(94.2ppm), respectively (**Figure 4**). These chemical shifts were largely consistent with literature values looking at HP [2-^13^C]pyruvate metabolism in human and rat brains ^28,29^. In comparison, the chemical shifts found in references (^28, 29^) for [5-^13^C]glutamate was estimated at (182.1, 182.1) ppm, [1-^13^C]ALCAR at (not-detected, 173.7) ppm, [2-^13^C]lactate centered at (70.1, 68.7) ppm, and [2-^13^C]pyruvate hydrate at (95.4, 94.6) ppm, respectively. The subtle, sub-ppm chemical shift discrepancy between our measured and literature values could be attributed to different compartments such as tissue, blood, intracellular vs extracellular, and microenvironment factors such as pH. Moreover, the putative assignments were also consistent with the known multiplicity profiles of these resonances, namely [5-^13^C]glutamate, [1-^13^C]ALCAR and [2-^13^C]hydrate as singlets, and [2-^13^C]lactate and [1,2-^13^C]pyruvate as doublets. The substrate and product identities can be further validated by their kinetic profiles (**Figures 5** & **6**). Pyruvate signal reached maximum at an earlier 12±2 s, whereas lactate, glutamate and ALCAR peaked later at 28±4 s, 30±3 s, and 28±4 s, respectively. These timings were calculated with a moving average of three timepoints to reduce temporal signal fluctuations.

**Figure 4.**
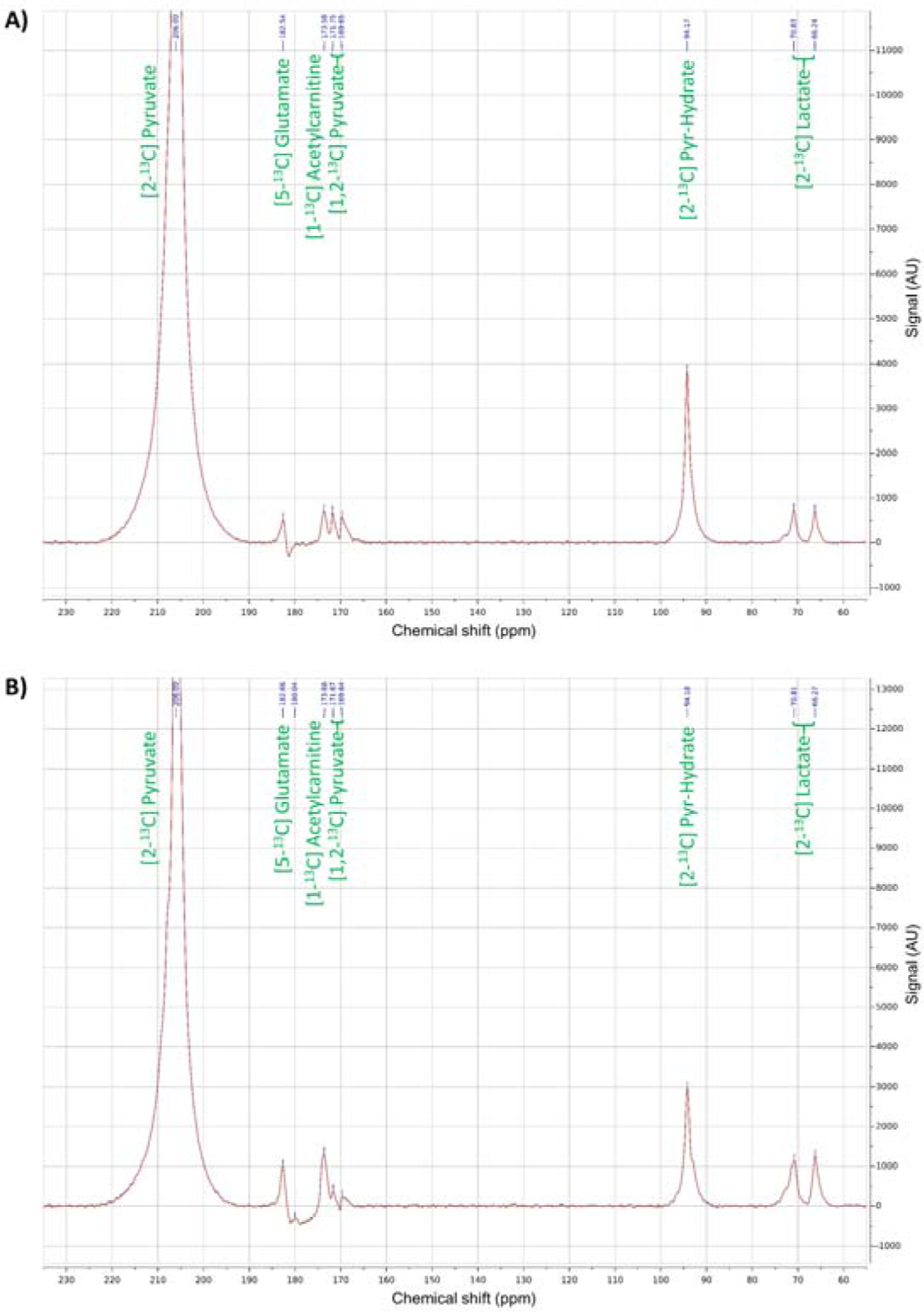
Representative summed **A)** fasting and **B)** fed spectra (Subject 1) reflecting TCA cycle energetics in human heart interrogated using hyperpolarized C2 pyruvate. The substrates and products in the spectra were assigned [5-^13^C]glutamate(182.5ppm), [1-^13^C]acetylcarnitine(ALCAR, 173.6ppm), [2-^13^C]lactate(70.8, 66.2ppm), [1,2-^13^C]pyruvate(171.8, 169.7ppm), and [2-^13^C]pyruvate hydrate(94.2ppm), respectively.

**Figure 5.**
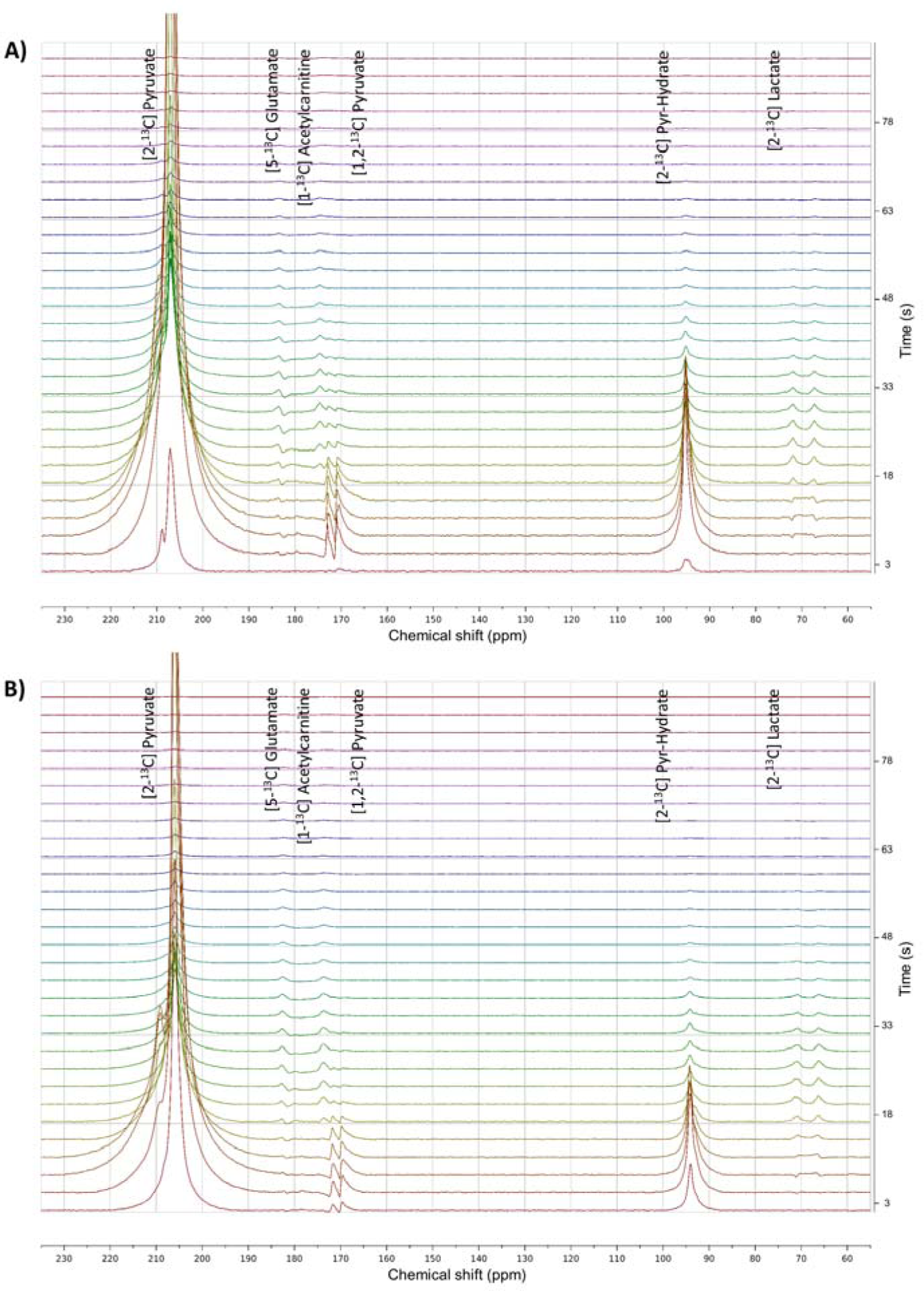
Representative dynamic **A)** fasting and **B)** fed spectra over time (Subject 1) showing metabolism of C2 pyruvate in human heart at fasting and fed states. The substrates ([2-^13^C]pyruvate, [1,2-^13^C]pyruvate, [2-^13^C]pyruvate-hydrate) and products ([5-^13^C]glutamate, [2-^13^C]lactate, [1-^13^C]acetylcarnitine) assignment was consistent with their kinetic profiles. The substrate reached maximum around 12s, whereas the products peaked later at approximately 28-30s. The spectral SNR was enhanced with a simple SVD filter as previously described^44^.

**Figure 6.**
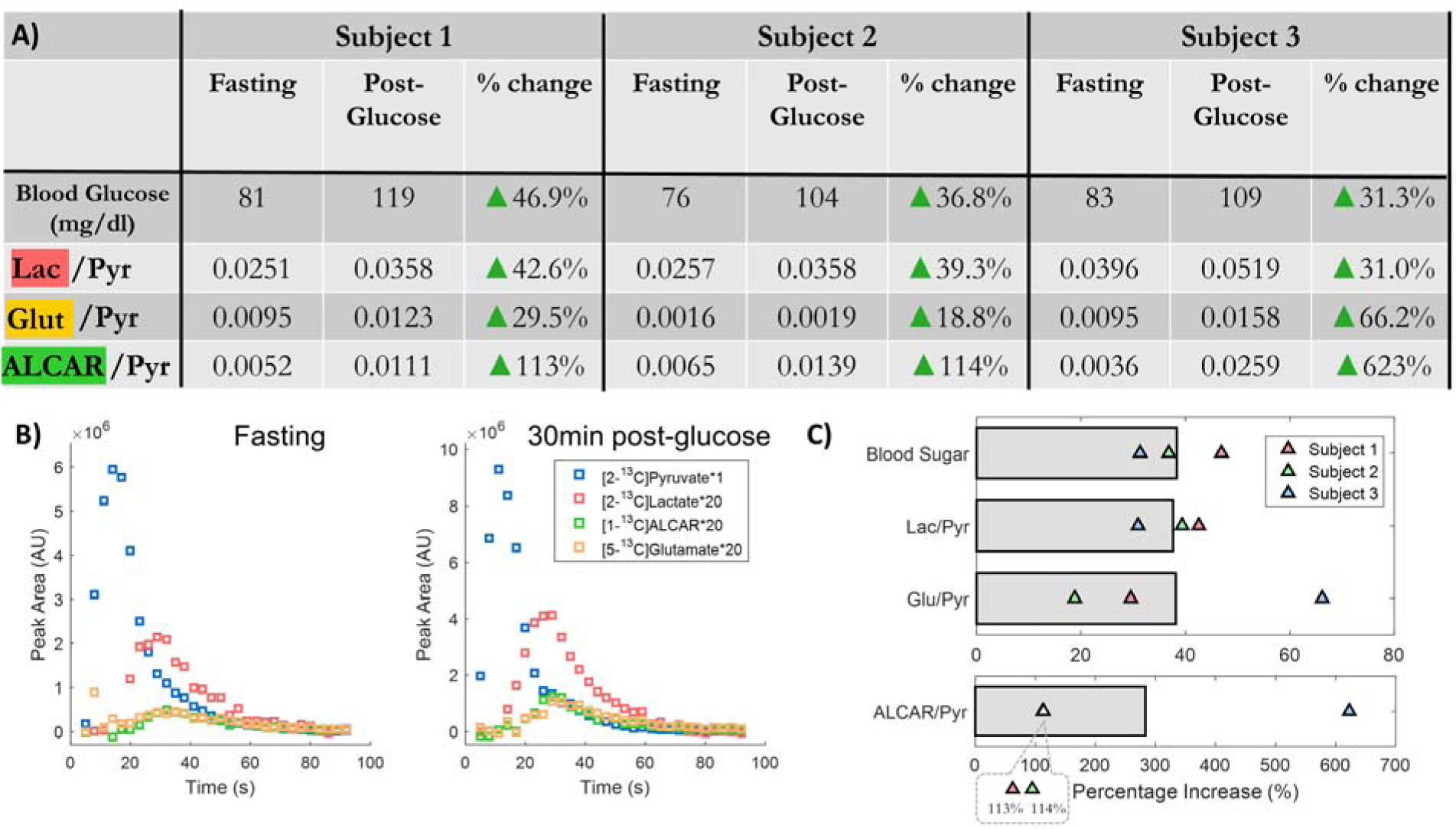
A) Summary of the changes of blood glucose levels and the metabolite/pyruvate ratios for the three study subjects at baseline fasting and 30-minutes after glucose load. **B)** Temporal evolution of the injected [2-^13^C]pyruvate and downstream metabolites [2-^13^C]lactate, [5-^13^C]glutamate and [1-^13^C]acetylcarnitine(ALCAR) in Subject 1 at baseline fasting and 30-minutes after oral glucose load. Time was calculated from the start of injection. **C)** Graphical representation of percentage increase after glucose load versus fasting. The bars indicate mean values for the three subjects.

The AUC ratio between [2-^13^C]pyruvate and [1,2-^13^C]pyruvate was approximately 50 fold, agreeing with the natural abundance of ^13^C isotope at the unenriched C1 position (notwithstanding possible differences in polarization levels and T_1_ relaxation times).

Median apparent SNRs of the metabolic species of interest were 910 for [2-^13^C]pyruvate, 25.8 for [2-^13^C]lactate, 6.20 for [5-^13^C]glutamate, and 5.92 for [1-^13^C]ALCAR in the fasting scan. SNR values for each fasting study was listed in **Supplemental Table 1**.

### Oral Glucose Load Test

The blood glucose readings for each subject are summarized in **Figure 6A**. Briefly, blood glucose levels were 76-83 mg/dl (median: 81 mg/dl) during fasting, and 104-119 mg/dl (median: 109 mg/dl) 30 minutes after intake of a sugar drink. The mean glucose level increase was 31 mg/dl, or 38.3% among the study subjects.

The metabolic product-to-pyruvate ratios before and after glucose load are summarized in **Figure 6A**. Overall, normalized levels of glycolytic and TCA-associated intermediates including lactate, glutamate, and ALCAR all increased after oral glucose administration. Among the three volunteers, there was 31.0-42.6% (median: 39.3%) increase of [2-^13^C]lactate, 18.8-66.2% (median: 29.5%) increase of [5-^13^C]glutamate, and 113-623% (median: 114%) increase of [1-^13^C]ALCAR with respect to substrate [2-^13^C]pyruvate measurements. In comparison, a prior study reported a more modest 15% (statistically nonsignificant) increase of [1-^13^C]lactate after glucose load, detected via [1-^13^C]pyruvate^22^. Interestingly, the distribution of fasting metabolism appeared to be more variable for glutamate (σ/μ=0.66) than for lactate (0.27) or ALCAR (0.28), albeit without statistical power to calculate significance from this small cohort. The HP species and their ppm shifts were unchanged between fasting and fed states, and no additional chemical species were identified on the HP spectra after oral glucose load.

## 4. Discussion

In summary, we developed methods, performed the first-in-human HP [2-^13^C]pyruvate cardiac MRI, and assessed metabolic flexibility by examining ^13^C-label incorporation in the glycolytic derivative [2-^13^C]lactate, TCA-associated metabolite [5-^13^C]glutamate and [1-^13^C]acetylcarnitine, in the fasting/fed states.. The slice-selective spectroscopy approach provided adequate SNR for identification of substrate and products, and for quantitative analysis in this pilot study. Assignment of the metabolic products is in agreement with both the ppm shifts reported in the prior [2-^13^C]pyruvate human brain and animal studies ^28–30^, and the putative mechanisms whereby the human heart utilizes glucose and fatty acids as energy sources^3,48,49^. Although neither a direct chemical analysis nor isotope tracing of these cardiac metabolites would be feasible in humans, the agreement between mechanistic premises and experimental results provided sufficiently strong evidence as to the identities of the observed products (i.e. [2-^13^C]lactate, [5-^13^C]glutamate and [1-^13^C]acetylcarnitine).

Pyruvate isotopically labelled at the C2 position allows the labelled carbon to enter the TCA cycle, as opposed to a C1 label that comes off during the preceding decarboxylation step. [2-^13^C]pyruvate offers the unique capability to directly observe and quantify TCA cycle modulations in real-time that are inaccessible through [1-^13^C]pyruvate (**Figure 1**). It is expected that HP [2-^13^C]pyruvate isotopic flux to the TCA-associated metabolite [5-^13^C]glutamate is influenced by PDH flux, alpha-ketoglutarate dehydrogenase activity, and TCA cycle fluxes. Isotopic flux to [1-^13^C]ALCAR is also influenced by PDH, but requires CAT activity and may also provide a readout of the carnitine shuttle and substrate buffering.

This is the first study to detect [1-^13^C]ALCAR in humans, as it was not detected in the prior study imaging HP [2-^13^C]pyruvate in the healthy human brain^28^ – this is not unexpected, because CAT is highly expressed only in cardiac and skeletal muscles. On the other hand, the resonance at 179.6ppm identified in the brain, putatively assigned [5-^13^C]glutamine and/or [1-^13^C]citrate, was not detected in the heart. Prior rodent heart HP [2-^13^C]pyruvate studies reported detection of [1-^13^C]citrate^12,30^, but at lower levels than [2-^13^C]lactate, [5-^13^C]glutamate and [1-^13^C]acetylcarnitine, suggesting [1-^13^C]citrate was below the detection limit in these studies.

The ability to simultaneously measure [5-^13^C]glutamate and [1-^13^C]acetylcarnitine provides much more specific measurements of mitochondrial energy metabolism when compared with just PDH flux (e.g. [1-^13^C]pyruvate to ^13^C-bicarbonate). HP [2-^13^C]pyruvate to [5-^13^C]glutamate requires PDH and TCA cycle flux. Meanwhile, HP [2-^13^C]pyruvate to [1-^13^C]acetylcarnitine requires PDH flux to acetyl-CoA, but then it does not enter the TCA cycle and rather is converted to acetylcarnitine via CAT, which is highly expressed in cardiac myocytes. Conversion of acetyl-CoA to acetylcarnitine as opposed to TCA cycle flux could be a result of excess acetyl-CoA, where prior studies have suggested this conversion acts as a buffer^12^ to fine tune acetyl-CoA availability (akin to phosphocreatine that serves as an ATP buffer). The Randle hypothesis also predicts that acetyl-CoA molecules derived from glucose and FA oxidation compete for entry into the TCA cycle^48^, implying that [2-^13^C]pyruvate to [1-^13^C]acetylcarnitine production could be modulated based on the presence of excess acetyl-CoA from either glucose or FA oxidation. The acetylcarnitine can then be transported into the cytosol for reuse. Acetylcarnitine influences protein function and gene expression by serving as an acetyl donor^50^ for acetylation of mitochondrial/cytosolic proteins and histones. Importantly, tissue levels of acetylcarnitine can be increased by oral supplementation^51^ with L-carnitine or acetylcarnitine^52^, thus providing an avenue for influencing mitochondrial function, myocardial flexibility and energetics.

Our results in response to the oral glucose load showed increases in both [5-^13^C]glutamate and [1-^13^C]acetylcarnitine in all three subjects, suggesting increased PDH flux and glucose oxidation in the fed state as expected by the Randle cycle ^31,53^. However, there were notably much larger increases in [1-^13^C]acetylcarnitine compared to [5-^13^C]glutamate (normalized to substrate [2-^13^C]pyruvate, **Figure 6**). This could be due to an excess of acetyl-CoA from glucose oxidation after the glucose load that is being buffered via CAT to acetylcarnitine. This would mean that just capturing PDH flux using HP [1-^13^C]pyruvate to ^13^C-bicarbonate would not correlate with TCA cycle flux, as varying amounts of acetyl-CoA will be buffered into acetylcarnitine. Our data showing differential changes in [5-^13^C]glutamate and [1-^13^C]acetylcarnitine following oral glucose load indicate that PDH flux and TCA cycle flux are not coupled across fasted and fed states. Thus, we believe that HP [2-^13^C]pyruvate is superior to HP [1-^13^C]pyruvate for assessing measuring metabolism and mitochondrial function because it provides greater information on TCA cycle flux, which is not accessible with HP [1-^13^C]pyruvate.

### This pilot study looking at HP [2-^13^C]pyruvate in the human heart has a few limitations

First of all, the slice-selective spectroscopy approach limited the ability to spatially localize the substrate and product chemical species. To a certain extent, this was mitigated by a rudimentary localization provided by positioning the two ^13^C paddle receiver arrays over the heart. As seen in **Figure 2**, the region covered by the intersection of the coil sensitivity profile and the slice selective RF pulse contained the heart, lungs, the rib cage, and some cutaneous and subcutaneous tissues of the chest wall. Considering the total blood volume, signal would originate predominately from the heart, whereas contribution from the remainder anatomy would be relatively negligible. This was supported by findings from HP ^13^C cardiac imaging studies in the literature^21,45,46^. The main challenge of the spectroscopy approach would be to distinguish the signal originating from the myocardium versus the chamber cavities, where the former is of our primary interest. Imaging-based methods using either metabolite-specific acquisition or model-based chemical shift imaging can be developed to offer such capabilities in the future ^54–56^.

The three subjects happened to be all male. Future enrollment of female subjects in a larger cohort is needed to generalize these outcomes. Although there are some early suggestions that cardiac metabolism and cardiomyopathy presentation between different sexes^57,58^, experimental validation is still required to draw relevant conclusions.

The protocol did not include additional biomarker measurements, such as serum hemoglobin A1c (HbA1c), fatty acid levels, or a metabolic panel, that might be useful for interpretation of the metabolic imaging. For example, differences in HbA1c, reflecting relatively long-term glucose utilization^59,60^, may provide explanation for the inter-subject variation of both fasting metabolism detected via HP [2-^13^C]pyruvate, as well as differences in the magnitude of responses to glucose challenge.

The small number of subjects precludes assessment of the relationship between physical activity and cardiac acetylcarnitine levels, which are influenced by CAT levels/activity, carnitine levels as well as PDH flux and FA oxidation.

## 5. Conclusions

This investigation demonstrated for the first time, hyperpolarized [2-^13^C]pyruvate MRI measurements of metabolic intermediates of the TCA cycle and acetylcarnitine in the fasting and fed states in human heart. We developed an MR spectroscopic approach using a broadband, spectral-spatially selective RF excitation to observe HP [2-^13^C]pyruvate and its metabolic conversion to [2-^13^C]lactate, [5-^13^C]glutamate and [1-^13^C]ALCAR in vivo, in human myocardium. Compared to prior HP [1-^13^C]pyruvate studies, HP [2-^13^C]pyruvate provided novel detection of [5-^13^C]glutamate and [1-^13^C]acetylcarnitine that are not possible with HP [1-^13^C]pyruvate or other imaging techniques. The pharmaceutical and MR methods for HP [2-^13^C]pyruvate developed in this study enable future studies of metabolic flexibility in the human heart.

## Data Availability

The datasets analyzed during the current study are available from the corresponding author on reasonable request.

## Acknowledgements

We would like to thank Kiersten Cheung, Heather Daniels, Romelyn Delos Santos, Evelyn Escobar, Mary Frost, Dr. Philip Lee, and Kimberly Okamoto for their assistance with the patient studies. This work was supported by grants from the UCSF Resource Allocation Program Team Science Award, Myokardia Inc. Myoseeds program, NIH (R33HL161816, P41EB013598), and a postdoctoral fellowship from the American Heart Association.

## Abbreviations

TCA cycle: tricarboxylic acid cycle
FA: fatty acid
ALCAR: acetylcarnitine
CAT: carnitine acetyltransferase

---

**Supplemental Table 1.** The table summarizes the measured summed apparent SNR (aSNR) for pyruvate, lactate, glutamate and ALCAR at the baseline fasting scan. Overall, the aSNR was sufficient for quantitative analysis of cardiac metabolism.

| SNR (unitless) | Subject 1 | Subject 2 | Subject 3 |
| --- | --- | --- | --- |
| Pyruvate | 1420 | 910 | 652 |
| Lactate | 35.6 | 23.4 | 25.8 |
| Glutamate | 13.5 | 1.46 | 6.20 |
| ALCAR | 7.38 | 5.92 | 2.35 |

## References

1. Roth GA, Mensah GA, Johnson CO, et al. Global Burden of Cardiovascular Diseases and Risk Factors, 1990-2019: Update From the GBD 2019 Study. J Am Coll Cardiol. Dec 22 2020;76(25):2982–3021. doi:10.1016/j.jacc.2020.11.010

2. Kolwicz SC, Jr., Purohit S, Tian R. Cardiac metabolism and its interactions with contraction, growth, and survival of cardiomyocytes. Circ Res. Aug 16 2013;113(5):603–16. doi:10.1161/CIRCRESAHA.113.302095

3. Lopaschuk GD, Ussher JR, Folmes CD, Jaswal JS, Stanley WC. Myocardial fatty acid metabolism in health and disease. Physiol Rev. Jan 2010;90(1):207–58. doi:10.1152/physrev.00015.2009

4. Vakrou S, Abraham MR. Hypertrophic cardiomyopathy: a heart in need of an energy bar? Front Physiol. 2014;5:309. doi:10.3389/fphys.2014.00309

5. Taegtmeyer H. Tracing cardiac metabolism in vivo: one substrate at a time. Journal of Nuclear Medicine. 2010;51(Supplement 1):80S–87S.

6. Handley MG, Medina RA, Nagel E, Blower PJ, Southworth R. PET imaging of cardiac hypoxia: opportunities and challenges. J Mol Cell Cardiol. Nov 2011;51(5):640–50. doi:10.1016/j.yjmcc.2011.07.005

7. Lopaschuk GD, Kelly DP. Signalling in cardiac metabolism. Cardiovasc Res. Jul 15 2008;79(2):205–7. doi:10.1093/cvr/cvn134

8. Taegtmeyer H, Young ME, Lopaschuk GD, et al. Assessing Cardiac Metabolism: A Scientific Statement From the American Heart Association. Circ Res. May 13 2016;118(10):1659–701. doi:10.1161/RES.0000000000000097

9. Herrero P, Peterson LR, McGill JB, et al. Increased myocardial fatty acid metabolism in patients with type 1 diabetes mellitus. J Am Coll Cardiol. Feb 7 2006;47(3):598–604. doi:10.1016/j.jacc.2005.09.030

10. Mather KJ, DeGrado TR. Imaging of myocardial fatty acid oxidation. Biochim Biophys Acta. Oct 2016;1861(10):1535–43. doi:10.1016/j.bbalip.2016.02.019

11. Kurhanewicz J, Vigneron DB, Ardenkjaer-Larsen JH, et al. Hyperpolarized (13)C MRI: Path to Clinical Translation in Oncology. Neoplasia. Jan 2019;21(1):1–16. doi:10.1016/j.neo.2018.09.006

12. Schroeder MA, Atherton HJ, Dodd MS, et al. The cycling of acetyl-coenzyme A through acetylcarnitine buffers cardiac substrate supply: a hyperpolarized 13C magnetic resonance study. Circulation: Cardiovascular Imaging. 2012;5(2):201–209.

13. Wang T, Zhu XH, Li H, et al. Noninvasive assessment of myocardial energy metabolism and dynamics using in vivo deuterium MRS imaging. Magnetic resonance in medicine. Dec 2021;86(6):2899–2909. doi:10.1002/mrm.28914

14. De Feyter HM, Behar KL, Corbin ZA, et al. Deuterium metabolic imaging (DMI) for MRI-based 3D mapping of metabolism in vivo. Science advances. Aug 2018;4(8):eaat7314. doi:10.1126/sciadv.aat7314

15. Ardenkjaer-Larsen JH, Fridlund B, Gram A, et al. Increase in signal-to-noise ratio of > 10,000 times in liquid-state NMR. Proc Natl Acad Sci U S A. Sep 2 2003;100(18):10158–63. doi:10.1073/pnas.1733835100

16. Golman K, in ‘t Zandt R, Thaning M. Real-time metabolic imaging. Proc Natl Acad Sci U S A. Jul 25 2006;103(30):11270–5. doi:10.1073/pnas.0601319103

17. Wang ZJ, Ohliger MA, Larson PEZ, et al. Hyperpolarized (13)C MRI: State of the Art and Future Directions. Radiology. May 2019;291(2):273–284. doi:10.1148/radiol.2019182391

18. Nelson SJ, Kurhanewicz J, Vigneron DB, et al. Metabolic Imaging of Patients with Prostate Cancer Using Hyperpolarized [1-C-13]Pyruvate. Science Translational Medicine. Aug 14 2013;5(198)doi:ARTN 198ra108 10.1126/scitranslmed.3006070

19. Joergensen SH, Hansen ESS, Bogh N, et al. Detection of increased pyruvate dehydrogenase flux in the human heart during adenosine stress test using hyperpolarized [1-(13)C]pyruvate cardiovascular magnetic resonance imaging. J Cardiovasc Magn Reson. Jun 6 2022;24(1):34. doi:10.1186/s12968-022-00860-6

20. Qin H, Tang S, Riselli AM, et al. Clinical translation of hyperpolarized (13) C pyruvate and urea MRI for simultaneous metabolic and perfusion imaging. Magnetic resonance in medicine. Jan 2022;87(1):138–149. doi:10.1002/mrm.28965

21. Cunningham CH, Lau JY, Chen AP, et al. Hyperpolarized 13C Metabolic MRI of the Human Heart: Initial Experience. Circ Res. Nov 11 2016;119(11):1177–1182. doi:10.1161/CIRCRESAHA.116.309769

22. Rider OJ, Apps A, Miller J, et al. Noninvasive In Vivo Assessment of Cardiac Metabolism in the Healthy and Diabetic Human Heart Using Hyperpolarized (13)C MRI. Circ Res. Mar 13 2020;126(6):725–736. doi:10.1161/CIRCRESAHA.119.316260

23. Apps A, Lau JYC, Miller J, et al. Proof-of-Principle Demonstration of Direct Metabolic Imaging Following Myocardial Infarction Using Hyperpolarized 13C CMR. JACC Cardiovasc Imaging. Jun 2021;14(6):1285–1288. doi:10.1016/j.jcmg.2020.12.023

24. Park JM, Reed GD, Liticker J, et al. Effect of Doxorubicin on Myocardial Bicarbonate Production From Pyruvate Dehydrogenase in Women With Breast Cancer. Circ Res. Dec 4 2020;127(12):1568–1570. doi:10.1161/CIRCRESAHA.120.317970

25. Munzel T, Gori T, Keaney JF, Jr., Maack C, Daiber A. Pathophysiological role of oxidative stress in systolic and diastolic heart failure and its therapeutic implications. Eur Heart J. Oct 7 2015;36(38):2555–64. doi:10.1093/eurheartj/ehv305

26. Bertero E, Maack C. Metabolic remodelling in heart failure. Nat Rev Cardiol. Aug 2018;15(8):457–470. doi:10.1038/s41569-018-0044-6

27. Bedi KC, Jr., Snyder NW, Brandimarto J, et al. Evidence for Intramyocardial Disruption of Lipid Metabolism and Increased Myocardial Ketone Utilization in Advanced Human Heart Failure. Circulation. Feb 23 2016;133(8):706–16. doi:10.1161/CIRCULATIONAHA.115.017545

28. Chung BT, Chen HY, Gordon J, et al. First hyperpolarized [2-(13)C]pyruvate MR studies of human brain metabolism. J Magn Reson. Dec 2019;309:106617. doi:10.1016/j.jmr.2019.106617

29. Park JM, Josan S, Grafendorfer T, et al. Measuring mitochondrial metabolism in rat brain in vivo using MR Spectroscopy of hyperpolarized [2-(1)(3)C]pyruvate. NMR Biomed. Oct 2013;26(10):1197–203. doi:10.1002/nbm.2935

30. Josan S, Park JM, Hurd R, et al. In vivo investigation of cardiac metabolism in the rat using MRS of hyperpolarized [1-13C] and [2-13C]pyruvate. NMR Biomed. Dec 2013;26(12):1680–7. doi:10.1002/nbm.3003

31. Fillmore N, Mori J, Lopaschuk GD. Mitochondrial fatty acid oxidation alterations in heart failure, ischaemic heart disease and diabetic cardiomyopathy. Br J Pharmacol. Apr 2014;171(8):2080–90. doi:10.1111/bph.12475

32. Knottnerus SJG, Bleeker JC, Wust RCI, et al. Disorders of mitochondrial long-chain fatty acid oxidation and the carnitine shuttle. Rev Endocr Metab Disord. Mar 2018;19(1):93–106. doi:10.1007/s11154-018-9448-1

33. Pearson DJ, Tubbs PK. Carnitine and derivatives in rat tissues. Biochem J. Dec 1967;105(3):953–63. doi:10.1042/bj1050953

34. Flori A, Liserani M, Frijia F, et al. Real-time cardiac metabolism assessed with hyperpolarized [1-(13) C]acetate in a large-animal model. Contrast media & molecular imaging. May-Jun 2015;10(3):194–202. doi:10.1002/cmmi.1618

35. Yoshihara H, Bastiaansen JA, Karlsson M, Lerche MH, Comment A, Schwitter J. Myocardial fatty acid metabolism probed with hyperpolarized [1-13C] octanoate. Journal of Cardiovascular Magnetic Resonance. 2015;17(1):1–2.

36. Ball DR, Rowlands B, Dodd MS, et al. Hyperpolarized butyrate: a metabolic probe of short chain fatty acid metabolism in the heart. Magnetic resonance in medicine. May 2014;71(5):1663–9. doi:10.1002/mrm.24849

37. Chen J, Singh TK, Al Nemri S, Zaidi M, Billingsley KL, Park JM. Hyperpolarized [1-13C] Acetyl-l-Carnitine Probes Tricarboxylic Acid Cycle Activity In Vivo. ACS sensors. 2023;

38. Taylor HL, Jacobs DR, Jr., Schucker B, Knudsen J, Leon AS, Debacker G. A questionnaire for the assessment of leisure time physical activities. J Chronic Dis. 1978;31(12):741–55. doi:10.1016/0021-9681(78)90058-9

39. Park I, Larson PEZ, Gordon JW, et al. Development of methods and feasibility of using hyperpolarized carbon-13 imaging data for evaluating brain metabolism in patient studies. Magnetic resonance in medicine. Sep 2018;80(3):864–873. doi:10.1002/mrm.27077

40. American Diabetes Association Professional Practice C. 2. Classification and Diagnosis of Diabetes: Standards of Medical Care in Diabetes-2022. Diabetes Care. Jan 1 2022;45(Suppl 1):S17–S38. doi:10.2337/dc22-S002

41. Meyer CH, Pauly JM, Macovski A, Nishimura DG. Simultaneous spatial and spectral selective excitation. Magnetic resonance in medicine. Aug 1990;15(2):287–304. doi:10.1002/mrm.1910150211

42. Crane JC, Gordon JW, Chen HY, et al. Hyperpolarized 13C MRI data acquisition and analysis in prostate and brain at University of California, San Francisco. NMR in Biomedicine. 2021;34(5):e4280.

43. Grist JT, Hansen ESS, Sanchez-Heredia JD, et al. Creating a clinical platform for carbon-13 studies using the sodium-23 and proton resonances. Magnetic resonance in medicine. Oct 2020;84(4):1817–1827. doi:10.1002/mrm.28238

44. Chen HY, Autry AW, Brender JR, et al. Tensor image enhancement and optimal multichannel receiver combination analyses for human hyperpolarized (13) C MRSI. Magnetic resonance in medicine. Dec 2020;84(6):3351–3365. doi:10.1002/mrm.28328

45. Ma J, Chen J, Reed GD, et al. Cardiac T2 * measurement of hyperpolarized (13) C metabolites using metabolite-selective multi-echo spiral imaging. Magnetic resonance in medicine. Sep 2021;86(3):1494–1504. doi:10.1002/mrm.28796

46. Reed GD, Ma J, Park JM, et al. Characterization and compensation of f0 inhomogeneity artifact in spiral hyperpolarized (13) C imaging of the human heart. Magnetic resonance in medicine. Jul 2021;86(1):157–166. doi:10.1002/mrm.28691

47. Prevention CfDCa. Defining Adult Overweight & Obesity. 2022.

48. Muoio DM, Noland RC, Kovalik JP, et al. Muscle-specific deletion of carnitine acetyltransferase compromises glucose tolerance and metabolic flexibility. Cell Metab. May 2 2012;15(5):764–77. doi:10.1016/j.cmet.2012.04.005

49. Tran DH, Wang ZV. Glucose Metabolism in Cardiac Hypertrophy and Heart Failure. J Am Heart Assoc. Jun 18 2019;8(12):e012673. doi:10.1161/JAHA.119.012673

50. De Loof M, Renguet E, Ginion A, et al. Enhanced protein acetylation initiates fatty acid-mediated inhibition of cardiac glucose transport. Am J Physiol Heart Circ Physiol. Mar 1 2023;324(3):H305–H317. doi:10.1152/ajpheart.00449.2022

51. Tragni V, Primiano G, Tummolo A, et al. Personalized Medicine in Mitochondrial Health and Disease: Molecular Basis of Therapeutic Approaches Based on Nutritional Supplements and Their Analogs. Molecules. May 29 2022;27(11)doi:10.3390/molecules27113494

52. Kerner J, Yohannes E, Lee K, et al. Acetyl-L-carnitine increases mitochondrial protein acetylation in the aged rat heart. Mech Ageing Dev. Jan 2015;145:39–50. doi:10.1016/j.mad.2015.01.003

53. Heather LC, Clarke K. Metabolism, hypoxia and the diabetic heart. J Mol Cell Cardiol. Apr 2011;50(4):598–605. doi:10.1016/j.yjmcc.2011.01.007

54. Gordon JW, Chen HY, Dwork N, Tang S, Larson PEZ. Fast Imaging for Hyperpolarized MR Metabolic Imaging. J Magn Reson Imaging. Mar 2021;53(3):686–702. doi:10.1002/jmri.27070

55. Gordon JW, Vigneron DB, Larson PE. Development of a symmetric echo planar imaging framework for clinical translation of rapid dynamic hyperpolarized (13) C imaging. Magnetic resonance in medicine. Feb 2017;77(2):826–832. doi:10.1002/mrm.26123

56. Wiesinger F, Weidl E, Menzel MI, et al. IDEAL spiral CSI for dynamic metabolic MR imaging of hyperpolarized [1-13C]pyruvate. Magnetic resonance in medicine. Jul 2012;68(1):8–16. doi:10.1002/mrm.23212

57. Murphy E, Amanakis G, Fillmore N, Parks RJ, Sun J. Sex Differences in Metabolic Cardiomyopathy. Cardiovasc Res. Mar 15 2017;113(4):370–377. doi:10.1093/cvr/cvx008

58. Wittnich C, Tan L, Wallen J, Belanger M. Sex differences in myocardial metabolism and cardiac function: an emerging concept. Pflugers Arch. May 2013;465(5):719–29. doi:10.1007/s00424-013-1232-1

59. Singer DE, Nathan DM, Anderson KM, Wilson PW, Evans JC. Association of HbA1c with prevalent cardiovascular disease in the original cohort of the Framingham Heart Study. Diabetes. Feb 1992;41(2):202–8. doi:10.2337/diab.41.2.202

60. Cavero-Redondo I, Peleteiro B, Álvarez-Bueno C, Rodriguez-Artalejo F, Martínez-Vizcaíno V. Glycated haemoglobin A1c as a risk factor of cardiovascular outcomes and all-cause mortality in diabetic and non-diabetic populations: a systematic review and meta-analysis. BMJ open. 2017;7(7):e015949.

